# Advancing Healthcare AI Governance: A Comprehensive Maturity Model Based on Systematic Review

**DOI:** 10.1101/2024.12.30.24319785

**Authors:** Rowan Hussein, Anna Zink, Bashar Ramadan, Frederick M Howard, Maia Hightower, Sachin Shah, Brett K Beaulieu-Jones

**Affiliations:** Center for Computational Medicine and Clinical Artificial Intelligence, Department of Medicine, University of Chicago; Center for Applied AI, Booth School of Business, University of Chicago; Equality AI, Park City, UT

## Abstract

Artificial Intelligence (AI) deployment in healthcare is accelerating, yet comprehensive governance frameworks remain fragmented and often assume extensive resources. Through a systematic review of 22 frameworks published between 2019-2024, we identified seven critical domains of healthcare AI governance: organizational structure, problem formulation, external product evaluation, algorithm development, model evaluation, deployment integration, and monitoring maintenance. While existing frameworks provide valuable guidance, they frequently target only large academic medical centers, creating barriers for smaller healthcare organizations. To address this gap, we propose the Healthcare AI governance Readiness Assessment (HAIRA), a five-level maturity model that provides actionable governance pathways based on organizational resources and capabilities. HAIRA spans from Level 1 (Initial / Ad Hoc) suitable for small practices to Level 5 (Leading) for major academic centers, with specific benchmarks across all seven governance domains. This tiered approach enables healthcare organizations to assess their current AI governance capabilities and establish appropriate advancement targets. Our framework addresses a critical need for adaptive governance strategies that can support AI-enabled healthcare value across diverse settings and ensures that AI implementation delivers tangible benefits to healthcare systems of varying sizes and resource levels.

## Introduction

Artificial Intelligence (AI) is being rapidly deployed in many aspects of healthcare with both predictive and generative AI applications poised for continued expansion^1^. These technologies show considerable promise in addressing critical healthcare challenges, such as offering clinical decision support based on predictive models of patient risk or streamlining administrative tasks for physicians^2,3^. The U.S. Food and Drug Administration has proposed a regulatory framework for AI/ML software serving as a medical device^4^, and the European Union (EU) AI Act^5^ creates similar safeguards for applications for EU member states. Successful governance of new technologies in healthcare requires a structured approach to manage their implementation as well as to foresee, measure, and mitigate the consequences of the new technology. This is particularly crucial in medical settings, where suboptimal performance or unexpected outcomes can have life-altering implications. For instance, inadequately governed AI tools may inadvertently perpetuate systemic inequities, as evidenced by algorithms that unintentionally deprioritize care for underserved populations^6^. Effective governance is essential to ensure that technological advancements in healthcare are ethically implemented, efficiently utilized, and consistently prioritize patient safety.

As AI solutions have emerged, a wide variety of governance best practices in the form of frameworks, governance structures, checklists, and guidelines have been proposed.^7–23^ Many focus on ethical AI principles and methods for assessing the clinical need for AI tools algorithm development, implementation, and maintenance. However, there remains sparsity around recommendations for organizational structures, external product (e.g., models or integrated tools) evaluation, and the effects of AI implementation on downstream values and outcomes as opposed to intermediate metrics such as model accuracy. Some frameworks or aspects of frameworks are not feasible for many health systems due to resource and expertise constraints. Additionally, there is substantial variation between published recommendations, and it can be difficult to decide which approach fits each institution. These limitations illustrate the need for cohesive and comprehensive guidance to ensure responsible, successful, and effective AI/ML adoption in healthcare. Mature AI governance does not need to slow down the adoption of AI but should instead provide pathways to systematically identify and mitigate risks and weaknesses, accelerate the adoption among clinicians or other end users, compare between AI-based vendors, products, or models, deploy and integrate into workflows, measure its impact, and monitor for future performance degradation.

This study aimed to identify, analyze, and unify existing frameworks, governance structures, and checklists related to AI implementation in the healthcare sector. We evaluated these documents based on their outlined recommendations across key domains of the AI governance: organizational structure, problem formulation, e algorithm evaluation and selection, algorithm development and training, model evaluation and validation, workflow deployment and integration, and monitoring and maintenance. Existing proposed frameworks tended to be geared towards large academic health systems with substantial resources and personnel with expertise in many areas including but not limited to AI, data science, statistics, implementation science, healthcare IT and EMR integration, business intelligence and reporting, and quality improvement. There are few actionable pathways tailored to healthcare organizations with varying levels of resources and expertise. In synthesizing the overarching recommendations, we aimed to clarify existing gaps and provide guidance on the resources necessary to successfully adopt these recommendations. As a result, we propose a readiness evaluation framework entitled “Healthcare AI governance Readiness Assessment” (HAIRA). This framework aims to provide a method for healthcare systems to assess their current AI governance and to establish pragmatic targets based on availability of resources.

## Results

We first assessed the literature to understand the existing landscape for frameworks, checklists, and other forms of proposed best practices for the governance of AI in healthcare. To do this we conducted a search for articles proposing frameworks, guidelines, or checklists for artificial intelligence in healthcare (Figure 1, detailed in methods). Initially, the search of keywords generated 2,117 articles. Applying a temporal filter to focus on recent developments (2019-2024) reduced this to 1,182 articles. Further refinement by limiting the search to carefully selected relevant journals resulted in 137 articles. These articles were manually reviewed to identify those articles addressing or proposing structures for AI governance in health care. The search process resulted in a final inclusion of 22 articles for in-depth analysis.

**Figure 1.**
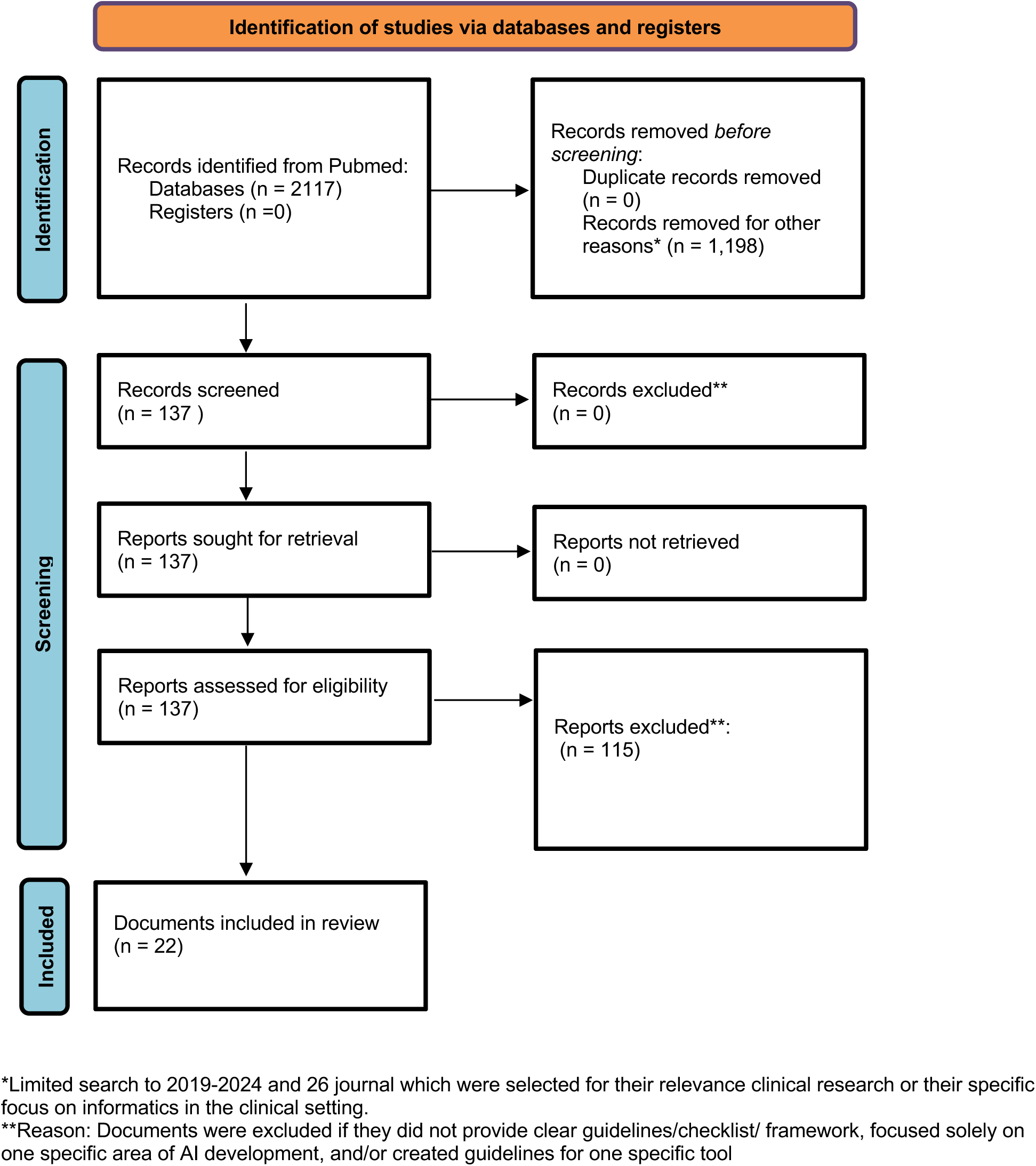
Delineates the systematic process for article identification and selection. Our initial search strategy focused on frameworks, governance structures, and checklists about artificial intelligence (AI) in healthcare. The original group of 2,117 articles was refined to 137 by applying two criteria: limiting the scope to 26 selected journals and constraining the publication timeframe to 2019-2024. After a comprehensive review, articles were excluded if they did not present clear guidelines or if they focused exclusively on a single AI model or specific aspect of AI implementation.

We reviewed and identified recommendations from the 22 selected articles. Recommendations were organized into the following seven categories based on widely recognized processes involved in the development and implementation of AI governance^24–26^:

1. *Organizational Structure*: Leadership and teams responsible for AI tool selection and evaluation.
2. *Problem Formulation:* Assessment of the clinical issue addressed by the AI tool, including input and output specifications.
3. *Algorithm Development and Model Training*: Application development, including algorithm design and data collection/dataset selection.
4. *External Product Evaluation and Selection:* Pre-launch testing by external parties to assess generalizability beyond the training population.
5. *Model Evaluation and Validation*: Evaluation by healthcare systems to ensure applicability to specific populations and assess potential risks, errors, or gaps.
6. *Deployment and Integration*: Implementation of the software/tool into healthcare system workflows.
7. *Monitoring and Maintenance*: Post-launch assessments to evaluate tool reliability and success as defined by initial problem formulation.

The selected articles and their recommendation categories are presented in Table 1. Across the literature, emphasis is placed on problem formulation, algorithm development/model training, and monitoring/maintenance. Conversely, organizational structure and external product evaluation received comparatively less attention.

**Table 1.**
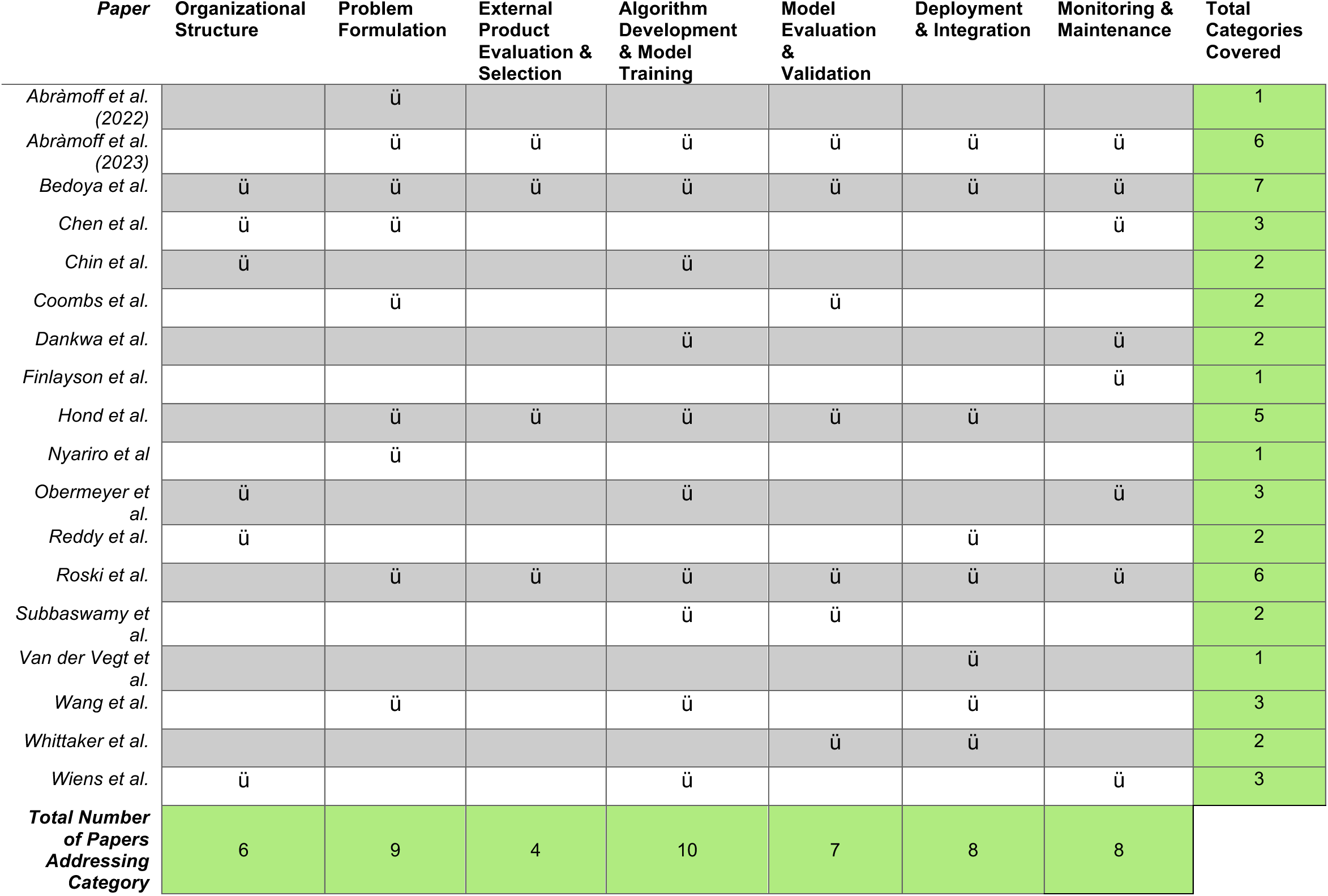
Compilation of documents with checklists, guidelines, and frameworks with a primary focus on the regulation of Artificial Intelligence (AI) implementation in the healthcare industry. Each document was reviewed, and recommendations were mapped to a specific domain of AI governance.

Six articles encompassed all or almost all of the seven domains of AI governance. The insights and recommendations from these comprehensive works have been analyzed and incorporated into our broader synthesis and generalizations. Below is a broad summary of their foci:

A) Abramoff et al. developed a framework focused on promoting health equity and bias throughout the AI lifecycle^7^. The framework is not intended to be a comprehensive strategy for mitigating all bias and risks but attempts to highlight critical areas of evaluation to ensure overall improvements. Many of the sentiments are echoed throughout recommendations in the other literature.
B) Aotearoa et al. designed a governance structure for the development and implementation of AI tools with a focus on the health services of New Zealand, which provided unique insight into designing guidance and tools tailored to a target population^22^. They considered the following domains: clinical leadership and decision-making, ethical principles in design, equity and inclusiveness, Maori engagement, consumer perspective, legal accountability, data guardianship, and technical methods. These domains differed slightly from those applied in most other guidelines due to their setting-specific tailoring. Although this type of checklist may not be generalizable it provides crucial insight on how guidelines can be tailored to a target population.
C) Bedoya et al. created a governance framework built around the AI/ML development and implementation process with several checkpoints, which act as software regulatory stop points^8^. This framework is unique as it has already been deployed in a healthcare system, the Duke University Health System, and at time of publication 52 models were regulating utilizing the framework.
D) Dagan et al. developed the OPTICA tool which emerged as the most detailed of all the frameworks evaluated and heavily touched all with great detail every aspects of the AI life cycle^27^. It is unique in its extensiveness as well as defining expertise required for different phases. However, similar to other models, there remains a question of which hospital systems can feasibly implement this guidance.
A) Roski et al.^18^ reviewed published literature to evaluate potential areas of risk around each phase of AI development and implementation and designed evidence-based practices for mitigations. However, it does not provide actionable guidelines in comparison to the other frameworks such as a checklist for implementation.
E) van de Vegt et al. combined five different frameworks including TRIPOD+AI to develop more actionable steps as opposed to recommendations centered around AI model, data pipeline, human-computer interface, and clinical workflow ^20^. This new framework was titled the SALIENT framework and consists of components and tasks a health system may need to conduct to complete tasks. This framework provides extensive and actionable guidance on developing the model, choosing and evaluating the data as well as how to implement it into clinical practice and how to monitor it once it has gone under development. Its stand-out feature is the focus on specific steps a healthcare system can take rather than overarching recommendations, but it does not discuss organizational structures.

As seen across comprehensive frameworks, a major question remains around the feasibility of all health systems to apply these steps.

### Review of Governance Recommendations

Table 2 consolidates key recommendations from existing frameworks, including those supported by at least two reviewed articles. By emphasizing recurring themes, it offers an initial framework for tackling governance challenges across the AI lifecycle.

**Table 2.** Harmonized recommendations based on inclusion in multiple published frameworks.

| Phase | Recommendations |
| --- | --- |
| **Organizational Structure** | • Create a governance committee consisting of diverse stakeholders including data scientist/ML learning experts, AI developers, clinical experts, ethical and legal experts, and patient/target group.<br>• Stakeholders should be involved in all aspects of AI governance including the deployment, monitoring, and general evaluation. |
| **Problem Formulation** | • Investigate the current standards of care to gain an understanding of the status quo, workflow, and context to develop a clear and objective rationale for use of AI.<br>• Outline specific objectives and outcomes for AI tools to achieve and articulate the motivation of choosing a specific model. |
| **External Product Evaluation** | • Governance panel or hospital system should evaluate data utilized for training to ensure it is representative and there is clear rationale. Conduct validation analysis to evaluate overall generalizability of data. |
| **Algorithm Development & Model Training** | • AI development team should confirm data is compliant with relevant privacy legislation and has taken measures to ensure privacy is maintained in the deployment and integration of the tool.<br>• Thorough assessment of collected data to confirm it is representative of the target population and aligned with the intended use, setting, and other relevant characteristics.<br>• Final structure should be well described and a committee should thoroughly evaluate how the tool will be implemented with a focus on workflow and UI design. |
| **Model Evaluation & Validation** | • Ensure the training data reflect the target population.<br>• Conduct bias evaluation and implement mitigation strategies |
| **Deployment & Integration** | • Educated end users on the objectives of the tool as well as expected outcomes, potential biases, legal framework, and other benefits and limitations. Evaluate for human biases as well.<br>• Create a system to identify potential sources of risks and where individuals can report errors, failures, and misses.<br>• Shadow deployment to assess for risks, errors, and biases ahead of full integration. |
| **Monitoring & Maintenance** | • A real-time system should be in place to log and audit and test bias, accuracy, predictability, and transparency of decision-making.<br>• Monitor for dataset shifts and feedback loops.<br>• Updates and adaptations should occur but cautiously and there should be measures to abort should there be major errors. |

#### Organizational Structure

A recurring theme across multiple articles was the necessity for an overarching governance body^7–10,17^. This entity would not only assist in selecting AI tools and software for implementation but also guide the validation, implementation, and monitoring processes. The literature generally recommended that this body comprise a diverse set of stakeholders, including data scientists and machine learning experts, AI developers versed in the technical aspects of the tools, clinical experts to provide insights on output accuracy and field-specific needs, ethical and legal experts, and representatives from patient and target groups. Emphasis was placed on integrating these experts throughout all phases of the AI lifecycle to ensure both efficacy and responsible implementation. Similar to governance of traditional clinical decision support tools - a larger governance structure will be needed for custom, i.e., internally developed AI tools as opposed to vendor, i.e., commercially developed solutions^28^.

Chen et al. and Reddy et al. proposed a structure of subpanels, overseen by an executive-level overarching committee, each with specific foci to spearhead different aspects of evaluation throughout the AI lifecycle^8,17^. For example, the monitoring phase could be led by a committee with IT expertise to ensure seamless integration of the model into clinical workflows and to evaluate potential errors. This multi-tiered approach aims to provide comprehensive oversight while allowing for specialized attention to each critical phase of AI implementation in healthcare settings.

While not formally described in the frameworks identified through the search, a recent trend has been the creation of Chief AI Officer or similar roles^29^. This trend mirrors the emergence of Chief Medical Informatics Officers in recent years, reflecting the need for specialized technical expertise to facilitate adoption and translation of technology in healthcare systems. As AI becomes increasingly integrated into clinical practice, healthcare organizations may need to adapt their leadership structures to empower an executive to dedicate a large portion or all of their effort toward developing the organization’s AI strategy, coordinating with governance committees and stakeholders to accelerate and ensure responsible adoption and oversight of AI in healthcare.

#### Problem Formulation

Most articles briefly addressed the importance of clear objectives for AI implementation in healthcare, with only a few offering actionable guidance^9,11,14,17,21^. Healthcare systems were advised to document and evaluate current standards of care to understand and discretize existing workflows. By contextualizing the status quo, health systems can realize both the need for specific AI tools as well as how these tools fit into their clinical workflow and align with population needs.

A subset of articles provided more detailed guidance. For instance, after identifying an aim, de Hond et al. suggested outlining clinical success criteria, for example specific patient outcomes, and assess potential risks, including prediction errors. This comprehensive approach aims to ensure that AI tools are designed or selected to meet specific healthcare needs align with quality and safety standards in clinical practice.

#### Algorithm Development and Model Training

Guidelines specific to algorithm development are crucial for ethical and complaint use of AI tools. They help to mitigate risk such as bias, privacy breaches and potential harm to patients while maintaining public trust in AI technology. An example denoted by Obermeyer et al. is that of an AI algorithm which utilized health care costs a proxy to health need leading to a false conclusion that Black patients were healthier than their white counterparts^6^. Errors such as these can have significant outcomes in the healthcare field emphasizing the need for regulations around algorithm development.

A thorough assessment of collected data is crucial to confirm its representativeness of the target population and alignment with the intended use and setting. The data should be sufficiently large to support generalizability, and all data processing steps should be documented for implementing systems. The tool’s structure, UI design, and software should be clearly outlined for evaluation committees to ensure workflow compatibility.

Regulatory bodies have recognized that the historical data serving as the foundation for AI model training has the potential to produce models that perpetuate bias present in routine clinical care. For example, the EU Artificial Intelligence Act mandates high-quality data for training, denotes the risk of bias through proxy variables, and suggests various strategies for mitigating bias in AI systems. Additionally, in response to feedback from their AI regulation plan, the FDA acknowledged the risk of bias in historical data and presented the following strategies: regulatory science efforts, emphasis on diverse patient populations, real-world performance monitoring, and transparency requirements for manufactures amongst other methods ^4^. A number of tools exist to assess and correct for bias present in training data or models, including AI Fairness 360^30^, Google Fairness Indicators^31^, Aequitas^32^, and EqualityAI^33^. Additionally, there have been several checklists for reporting details about clinical AI models in a way that allows others to critically evaluate them downstream^12,34^.

#### Model Evaluation, Validation, and External Product Evaluation

AI/ML models are characterized by the potential to ‘overfit’ on training data - identifying nonbiologic patterns that may predict the outcome of interest - and thus validation in independent populations is especially important. For example - in one commonly used public medical imaging dataset, 80% of cases with a pneumothorax had chest tubes in place. Models trained on this dataset may predict pneumothorax based on the presence of external devices (rather than the presence of a pneumothorax) - leading to false positive prediction for pneumothorax in a patient with an ECMO cannula^35^. Nonetheless - many approved devices are not externally validated^36^.

External product evaluation, selection, and model evaluation and validation were among the least discussed aspects in the reviewed literature, often with overlapping requirements. Despite this limited coverage, several key recommendations emerged. The primary suggestion across multiple sources was for external and internal validation dedicated to ensuring data representativeness and aligning quality and use with the intended application. The Office of National Coordinator for Health Information Technology’s (ONC) Health Data, Technology, and Interoperability (HTI-1) Final Rule mandates that EHR vendors disclose information about predictive decision support interventions, including AI, to its customers. Its section on algorithm transparency requires disclosure of training data representativeness, subpopulation performance, and pathways for bias evaluation and mitigation. This information can be used by AI governance committees as part of the external validation process and a framework for the types of information they may demand from other vendors.

Hond et al., Bedoya et al, and Coombs et al., emphasized the importance of using either retrospective or prospective data for validation and generalizability assessment. Overall, while specific methodologies varied, there was a consensus on the need for transparency in the evaluation process. For example, detailed documentation of evaluation criteria, methodologies, and results, making this information accessible to potential users - and such information will be required by regulatory bodies^7^.

#### Deployment and Integration

Deployment of AI tools in healthcare settings primarily focused on three critical areas: (i) shadow deployments, (ii) preemptive risk identification measures, and (iii) comprehensive end-user training.

To assess tool applicability and mitigate potential widespread consequences from errors, many sources recommended implementing a shadow deployment phase. During this period, health systems can evaluate risks, errors, and biases prior to full integration of the AI tool. This approach allows for real-world testing without compromising patient care, providing valuable insights into the tool’s performance in specific clinical contexts. Shadow deployments enable comparison of AI model performance with current operational standards and clinical outcomes, serving as a baseline for subsequent clinical evaluation protocols. Lastly, many sources recommended conducting a silent evaluation test or effectiveness assessment prior to deployment, emphasizing thorough testing in real-world settings before full implementation.

Ensuring proper AI system functionality and user acceptance requires a thorough understanding of workflow integration and potential failure modes. To effectively track these issues, healthcare institutions need to implement systems for identifying potential risk sources and establish clear channels for end-users to report errors. Some researchers suggest creating dedicated AI oversight committees to monitor and respond to these reports in real time. Care needs to be taken to ensure user interaction with AI tools remains meaningful and does not contribute to “alert fatigue^37^”.

A major area of concern and guidance was the education of end-users. There is a risk of both over-reliance on automated systems, which can lead to increased errors, as well as under-reliance, when end-users do not trust model outputs. To mitigate this, end-users should receive comprehensive education on the AI tool’s objectives, expected outcomes, and potential limitations or biases. This training should also address the users’ own biases, which may influence tool usage. Model interpretability can also prevent inappropriate reliance on AI models - in one study, when AI visually outlined suspicious regions on chest radiographs, radiologists were less likely to accept incorrect AI classifications^38^.

#### Monitoring and Maintenance

Frameworks largely emphasized the necessity of real-time monitoring for rapidly addressing major errors and implementing systems to audit and test bias, accuracy, and predictability. This ongoing assessment is essential for maintaining alignment between the AI system’s performance and the evolving needs of the target population.

Continuous assessment plays a vital role in confirming that the training data remains aligned with the characteristics of the target population. This process can effectively flag dataset shifts, which may occur when the distribution of training data significantly diverges from deployment data. Such shifts can manifest as changes in the distribution of input features, alterations in the relationship between input features and target variables, or modifications to the target variable itself. AI tools that learn from their outputs through feedback loops must be closely monitored, as unchecked adjustments can amplify errors or biases over time. Moreover, AI tools capable of learning from their own outputs, feedback loops, require scrutiny. Real-time monitoring provides a mechanism to assess whether these self-adjustments are valid or potentially contributing to errors. Most currently approved medical devices are based on ‘locked’ algorithms, where outputs do not change with continued use - but regulatory bodies will facilitate the assessment of more flexible models. For example, the FDA AI/ML framework requires specification of an algorithm change protocol (ACP) to define the key aspects of model retraining and performance evaluation for such continuously updating models^4^.

The dynamic nature of healthcare environments necessitates adaptations and updates to AI tools. Real-time monitoring serves as an early warning system, flagging when changes should be considered. However, implementing changes cautiously is crucial to avoid exacerbating risks. Additionally, continuous monitoring for data quality and model performance, with predefined thresholds for model retraining, ensures sustained effectiveness and adaptability of AI tools over time.

Furthermore, real-time monitoring facilitates the detection of potential biases that may emerge during deployment. By continuously analyzing the AI tool’s outputs across diverse patient populations, healthcare providers can identify and address any disparities in performance or recommendations that could lead to inequitable care. This ongoing evaluation is crucial for ensuring that AI tools maintain fairness and effectiveness across all demographic groups.

Additionally, regular audits of tool outputs and user interactions can identify any potential bias introduced by the end user.

#### Broad Themes and Challenges

Health equity was a significant theme across categories. Recommendations included incorporating diverse viewpoints into the teams that select and develop AI tools and evaluating health conditions and standards of care in the tool’s intended context to anticipate potential sources of bias that may arise. The impact of the AI tool on specific populations should be evaluated and monitored post-deployment. AI developers are encouraged to develop bias-identifying and mitigating tools.

A common challenge identified in these articles was balancing actionable recommendations with guidance applicable across diverse AI applications. For example, the TRIPOD + AI^12^ and MI-CLAIM^34^ checklists stood out for their comprehensive recommendations; however, they primarily focused on clinical prediction models and may not extend to newer generative AI applications, such as large language models in healthcare. This highlights the need for adaptable governance structures that can evolve standards and processes to address emerging technologies.

### Healthcare AI governance Readiness Assessment (HAIRA)

Given the combined guidance and recommendations across the included frameworks we propose a Healthcare AI governance Readiness Assessment (HAIRA) to provide actionable targets while addressing resource disparities (Table 3). This five-level maturity model recognizes that healthcare organizations vary significantly in their resources, expertise, and AI implementation needs. Drawing on the concepts of maturity seen in established frameworks like HIMSS’s digital transformation models, HAIRA provides a structured pathway for healthcare organizations to assess and advance their AI governance capabilities^39^. The model spans from Level 1 (Initial/Ad Hoc), suitable for small practices beginning to explore AI implementation, to Level 5 (Optimized), appropriate for leading academic health systems pioneering industry standards. Each level addresses the seven domains identified in the literature: organizational structure, problem formulation, external product evaluation, algorithm development, model evaluation, deployment integration, and monitoring maintenance. Supplemental table 2 provides a summary version of this information. By providing clear benchmarks across these domains, HAIRA enables healthcare organizations to set realistic governance goals aligned with their resources and clinical needs, while establishing clear direction for improvement.

**Table 3.**
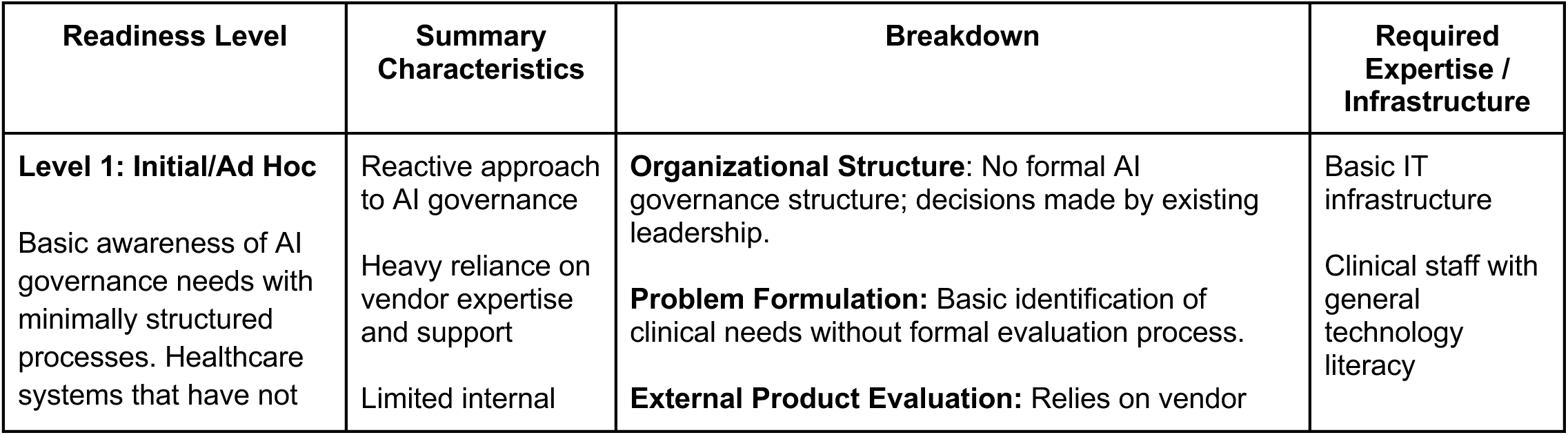
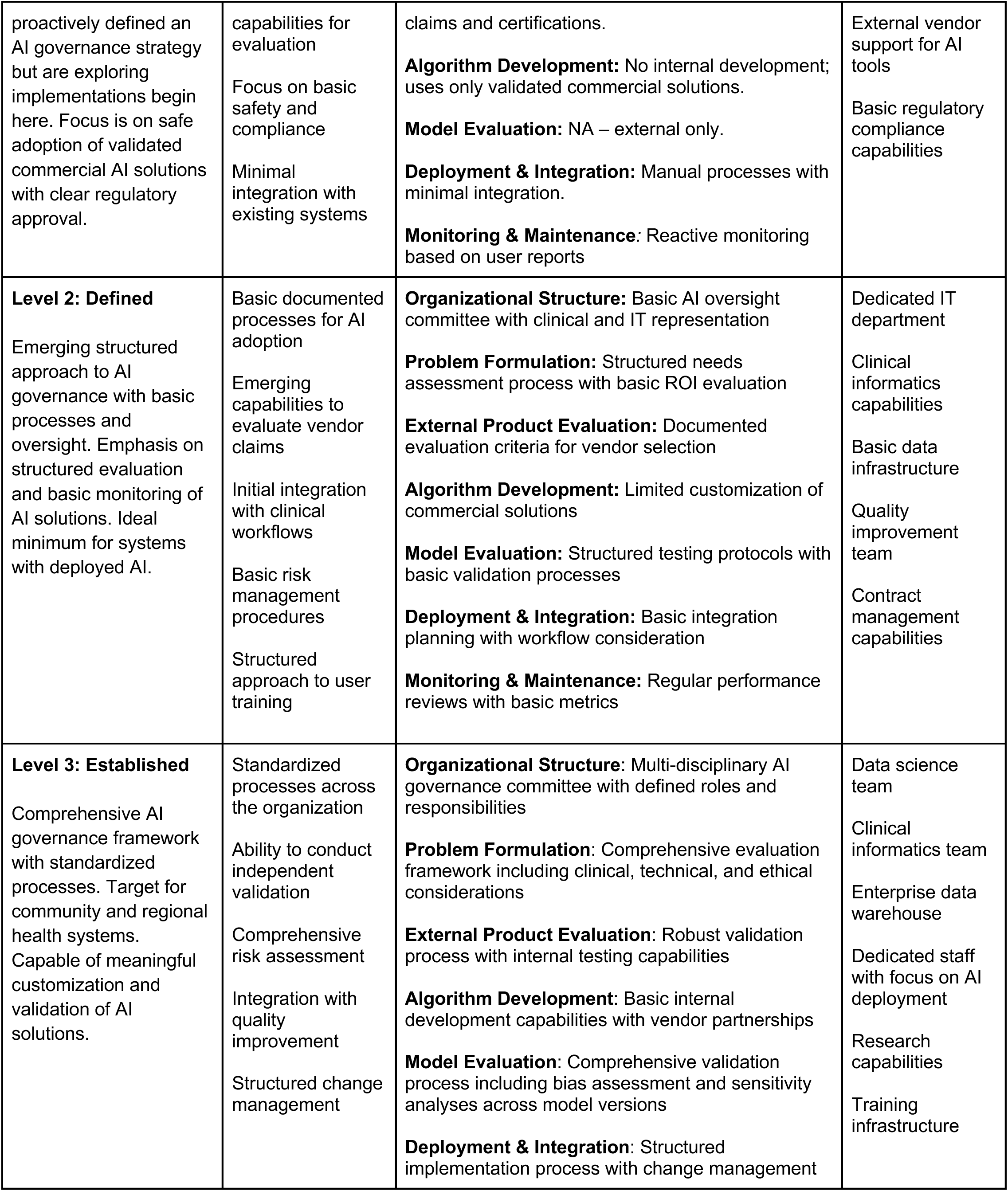
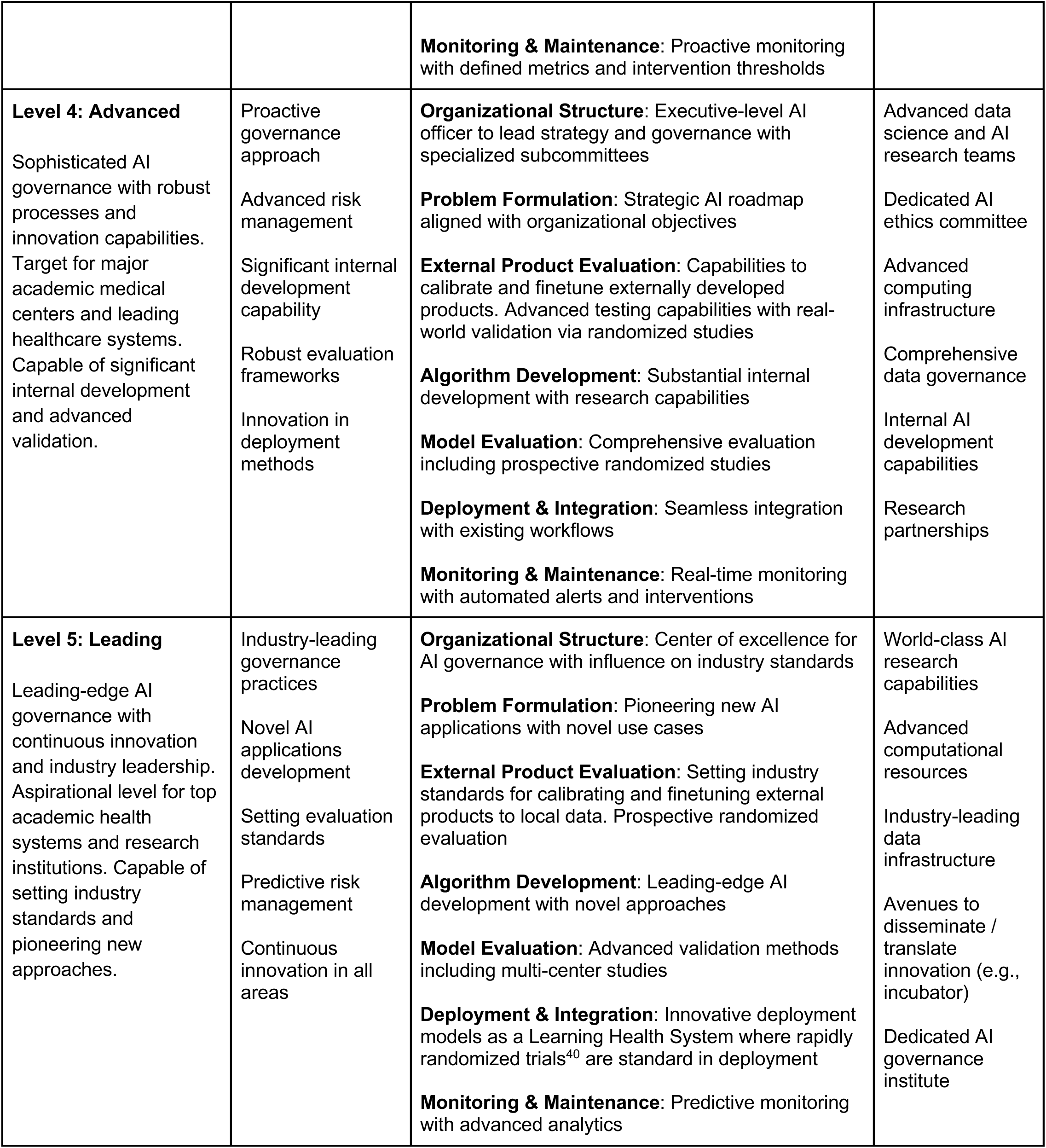
Healthcare AI governance Readiness Assessment (HAIRA).

| Readiness Level | Summary Characteristics | Breakdown | Required Expertise / Infrastructure |
| --- | --- | --- | --- |
| **Level 1: Initial/Ad Hoc**<br>Basic awareness of AI governance needs with minimally structured processes. Healthcare systems that have not | Reactive approach to AI governance<br>Heavy reliance on vendor expertise and support<br>Limited internal | **Organizational Structure:** No formal AI governance structure; decisions made by existing leadership.<br>**Problem Formulation:** Basic identification of clinical needs without formal evaluation process.<br>**External Product Evaluation:** Relies on vendor | Basic IT infrastructure<br>Clinical staff with general technology literacy |
| proactively defined an AI governance strategy but are exploring implementations begin here. Focus is on safe adoption of validated commercial AI solutions with clear regulatory approval. | capabilities for evaluation<br>Focus on basic safety and compliance<br>Minimal integration with existing systems | claims and certifications.<br>**Algorithm Development:** No internal development; uses only validated commercial solutions.<br>**Model Evaluation:** NA – external only.<br>**Deployment & Integration:** Manual processes with minimal integration.<br>**Monitoring & Maintenance:** Reactive monitoring based on user reports | External vendor support for AI tools<br>Basic regulatory compliance capabilities |
| **Level 2: Defined**<br>Emerging structured approach to AI governance with basic processes and oversight. Emphasis on structured evaluation and basic monitoring of AI solutions. Ideal minimum for systems with deployed AI. | Basic documented processes for AI adoption<br>Emerging capabilities to evaluate vendor claims<br>Initial integration with clinical workflows<br>Basic risk management procedures<br>Structured approach to user training | **Organizational Structure:** Basic AI oversight committee with clinical and IT representation<br>**Problem Formulation:** Structured needs assessment process with basic ROI evaluation<br>**External Product Evaluation:** Documented evaluation criteria for vendor selection<br>**Algorithm Development:** Limited customization of commercial solutions<br>**Model Evaluation:** Structured testing protocols with basic validation processes<br>**Deployment & Integration:** Basic integration planning with workflow consideration<br>**Monitoring & Maintenance:** Regular performance reviews with basic metrics | Dedicated IT department<br>Clinical informatics capabilities<br>Basic data infrastructure<br>Quality improvement team<br>Contract management capabilities |
| **Level 3: Established**<br>Comprehensive AI governance framework with standardized processes. Target for community and regional health systems. Capable of meaningful customization and validation of AI solutions. | Standardized processes across the organization<br>Ability to conduct independent validation<br>Comprehensive risk assessment<br>Integration with quality improvement<br>Structured change management | **Organizational Structure:** Multi-disciplinary AI governance committee with defined roles and responsibilities<br>**Problem Formulation:** Comprehensive evaluation framework including clinical, technical, and ethical considerations<br>**External Product Evaluation:** Robust validation process with internal testing capabilities<br>**Algorithm Development:** Basic internal development capabilities with vendor partnerships<br>**Model Evaluation:** Comprehensive validation process including bias assessment and sensitivity analyses across model versions<br>**Deployment & Integration:** Structured implementation process with change management | Data science team<br>Clinical informatics team<br>Enterprise data warehouse<br>Dedicated staff with focus on AI deployment<br>Research capabilities<br>Training infrastructure |
|  |  | **Monitoring & Maintenance:** Proactive monitoring with defined metrics and intervention thresholds |  |
| **Level 4: Advanced**<br>Sophisticated AI governance with robust processes and innovation capabilities. Target for major academic medical centers and leading healthcare systems. Capable of significant internal development and advanced validation. | Proactive governance approach<br>Advanced risk management<br>Significant internal development capability<br>Robust evaluation frameworks<br>Innovation in deployment methods | **Organizational Structure:** Executive-level AI officer to lead strategy and governance with specialized subcommittees<br>**Problem Formulation:** Strategic AI roadmap aligned with organizational objectives<br>**External Product Evaluation:** Capabilities to calibrate and finetune externally developed products. Advanced testing capabilities with real-world validation via randomized studies<br>**Algorithm Development:** Substantial internal development with research capabilities<br>**Model Evaluation:** Comprehensive evaluation including prospective randomized studies<br>**Deployment & Integration:** Seamless integration with existing workflows<br>**Monitoring & Maintenance:** Real-time monitoring with automated alerts and interventions | Advanced data science and AI research teams<br>Dedicated AI ethics committee<br>Advanced computing infrastructure<br>Comprehensive data governance<br>Internal AI development capabilities<br>Research partnerships |
| **Level 5: Leading**<br>Leading-edge AI governance with continuous innovation and industry leadership. Aspirational level for top academic health systems and research institutions. Capable of setting industry standards and pioneering new approaches. | Industry-leading governance practices<br>Novel AI applications development<br>Setting evaluation standards<br>Predictive risk management<br>Continuous innovation in all areas | **Organizational Structure:** Center of excellence for AI governance with influence on industry standards<br>**Problem Formulation:** Pioneering new AI applications with novel use cases<br>**External Product Evaluation:** Setting industry standards for calibrating and finetuning external products to local data. Prospective randomized evaluation<br>**Algorithm Development:** Leading-edge AI development with novel approaches<br>**Model Evaluation:** Advanced validation methods including multi-center studies<br>**Deployment & Integration:** Innovative deployment models as a Learning Health System where rapidly randomized trials<sup>40</sup> are standard in deployment<br>**Monitoring & Maintenance:** Predictive monitoring with advanced analytics | World-class AI research capabilities<br>Advanced computational resources<br>Industry-leading data infrastructure<br>Avenues to disseminate / translate innovation (e.g., incubator)<br>Dedicated AI governance institute |

The proposed system emphasizes adaptability, recognizing that not all healthcare systems have equal access to advanced infrastructure or specialized personnel. For example, lower-resource settings may focus initially on foundational steps such as data standardization and basic validation of vendor claims, while higher-resource systems can engage in more complex tasks like multimodal data integration or real-time monitoring.

## Discussion

This work identified both emerging and consistent themes of healthcare AI governance as well as significant discrepancies between frameworks. Many of the recommendations identified in individual articles highlight critical elements that should be considered by healthcare systems, underscoring the need for comprehensive and unified frameworks. However, the few frameworks that are comprehensive require substantial personnel, expertise, and resources — factors that may not be accessible to all healthcare systems.

In response to a need for actionable guidance that considers health system resources, we presented a new AI governance readiness model which we call HAIRA so that healthcare organizations can assess and advance their AI governance capabilities based on expert recommendations outlined in this review. Integrating this tiered system into existing implementation frameworks could help bridge gaps identified in current models. For instance, frameworks like SALIENT and OPTICA provide detailed guidance on specific stages of AI implementation but often assume access to advanced resources or expertise^20,27^. Many regional and community health systems do not have in-house data science capabilities but instead may have data analysts supporting traditional quality improvement and reporting activities. Health systems may even outsource substantial portions of their IT needs, including using a hosted EMR system. Most of the proposed governance frameworks would be exceedingly difficult to implement in these settings. By incorporating a tiered structure, these frameworks could offer actionable pathways for systems at varying levels of readiness, ensuring broader applicability.

While resource availability is a critical factor in implementing AI governance several other dimensions must also be considered. The demographic composition of the patient population significantly influences AI strategies, as diverse populations have distinct needs compared to homogeneous ones. Additionally, varying regulatory compliance and data privacy requirements necessitate customized governance approaches. Healthcare organizations should clearly define their objectives, as these goals inform the most suitable governance structures. For example, a system focused on enhancing diagnostic accuracy may require different oversight than one prioritizing operational efficiency. Finally, the existing organizational culture and established protocols play a crucial role in shaping AI governance implementation.

A significant challenge in evaluating AI governance frameworks in healthcare is the limited empirical evidence of their effectiveness. Many of these frameworks are relatively new, and their implementation has not been extensively tested or published. This lack of data makes it difficult to determine whether these governance structures lead to better outcomes or achieve their initial goals of mitigating risks and maintaining tool excellence. To address this gap, we recommend conducting randomized controlled trials (RCTs) to assess various frameworks and recommendations. These “rapid” and electronically driven RCTs have been shown to be feasible^40^, and could help identify which guidelines effectively support their intended objectives. Designing and executing such trials presents considerable challenges, given the complexity of healthcare environments and the rapid evolution of AI technologies but will be critical to accurately allocate and justify investments in clinical AI. Future research should focus on developing methodologies that go beyond retrospective analyses^41^ and instead conduct prospective randomized evaluations.

Ultimately, the development of adaptable frameworks tailored to different levels of healthcare system capability is critical for equitable AI adoption. Such frameworks would not only address the current gaps but also provide clear guidance for systems aiming to advance their technological capacity over time. This article contributes a more robust understanding of the current state and future directions of AI regulation in healthcare.

## Methods

Our assessment of the current landscape of AI regulatory practices in healthcare involved a multifaceted approach to ensure comprehensive coverage of relevant literature. We began by compiling a set of key terms related to AI regulation before refining them to generate a broad yet structured output. The final search string included articles with main subjects under the terms “Artificial Intelligence OR AI” AND “Health Care OR Healthcare,” AND “framework OR governance OR checklist.”

To further refine our search, we limited outputs to 26 journals (full list available in Supplementary Table 3). These journals were selected based on their broad relevance to the health field (e.g., *Nature Medicine*) or their specific focus on digital health, AI, or MI in healthcare contexts (e.g., *Lancet Digital Health*). Recognizing the rapid maturation of AI tools in healthcare, we restricted our search to publications from 2019-2024, ensuring the relevance and applicability of the regulatory frameworks examined.

From this curated list, we included reports and articles that focused on developing frameworks, checklists, or governance structures for regulating AI in healthcare settings. These documents were required to contain specific recommendations or guidelines. We prioritized articles addressing multiple aspects of the AI lifecycle. Based on expert feedback, we also incorporated articles that met our criteria but were not captured in the initial search.

## Data Availability

All data produced in the present work are contained in the manuscript

---

**Supplemental Table 1:**
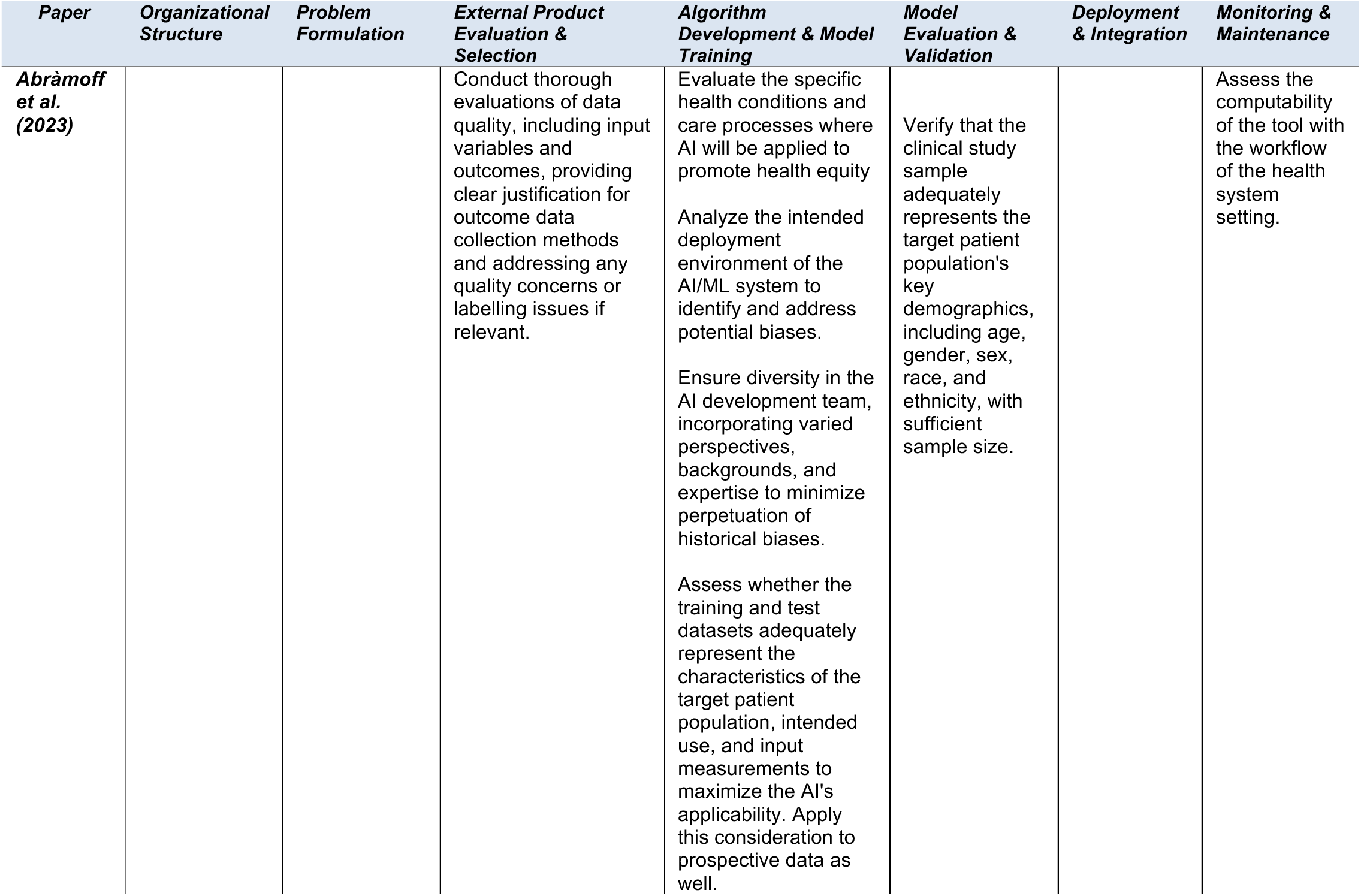
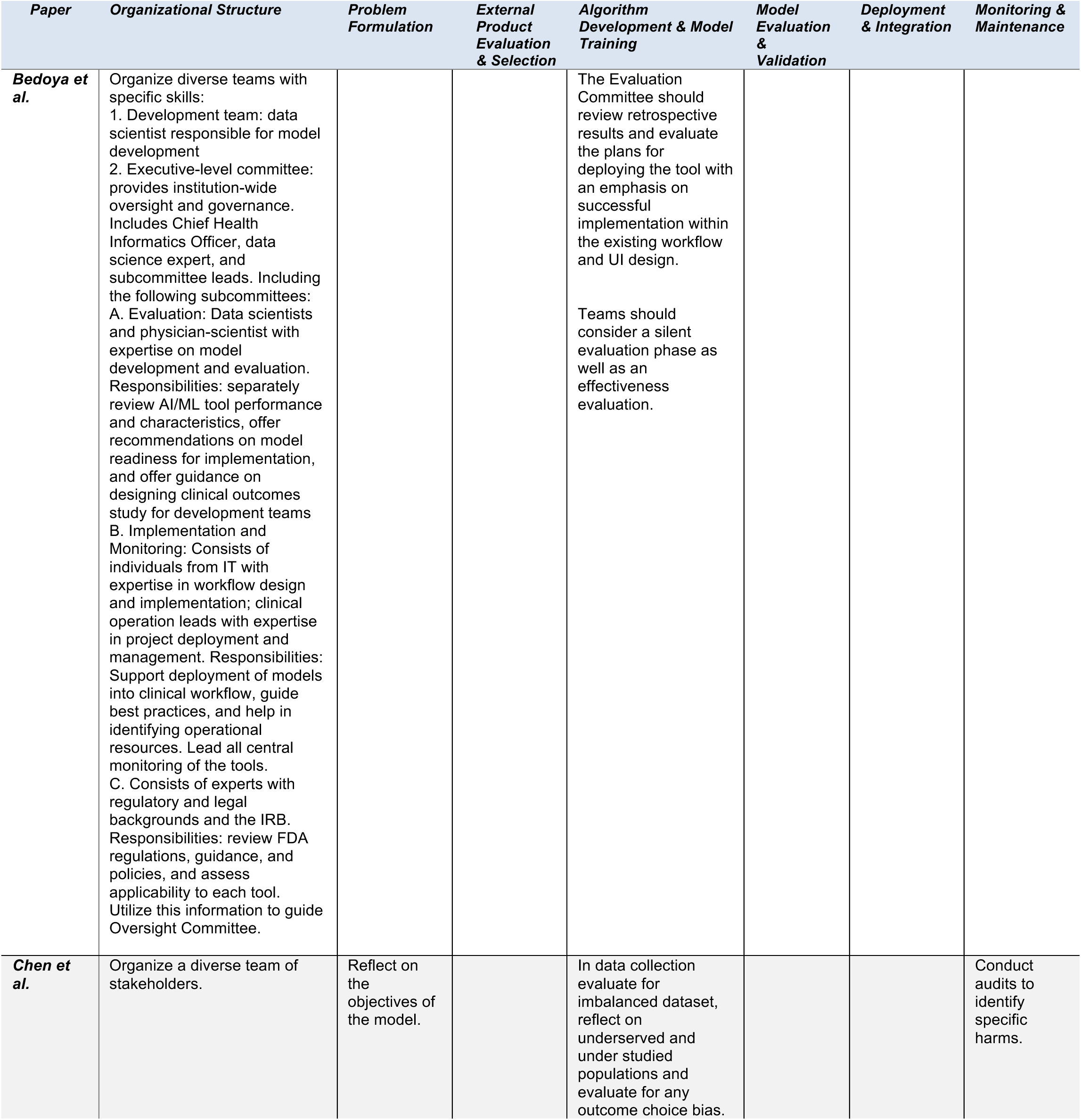
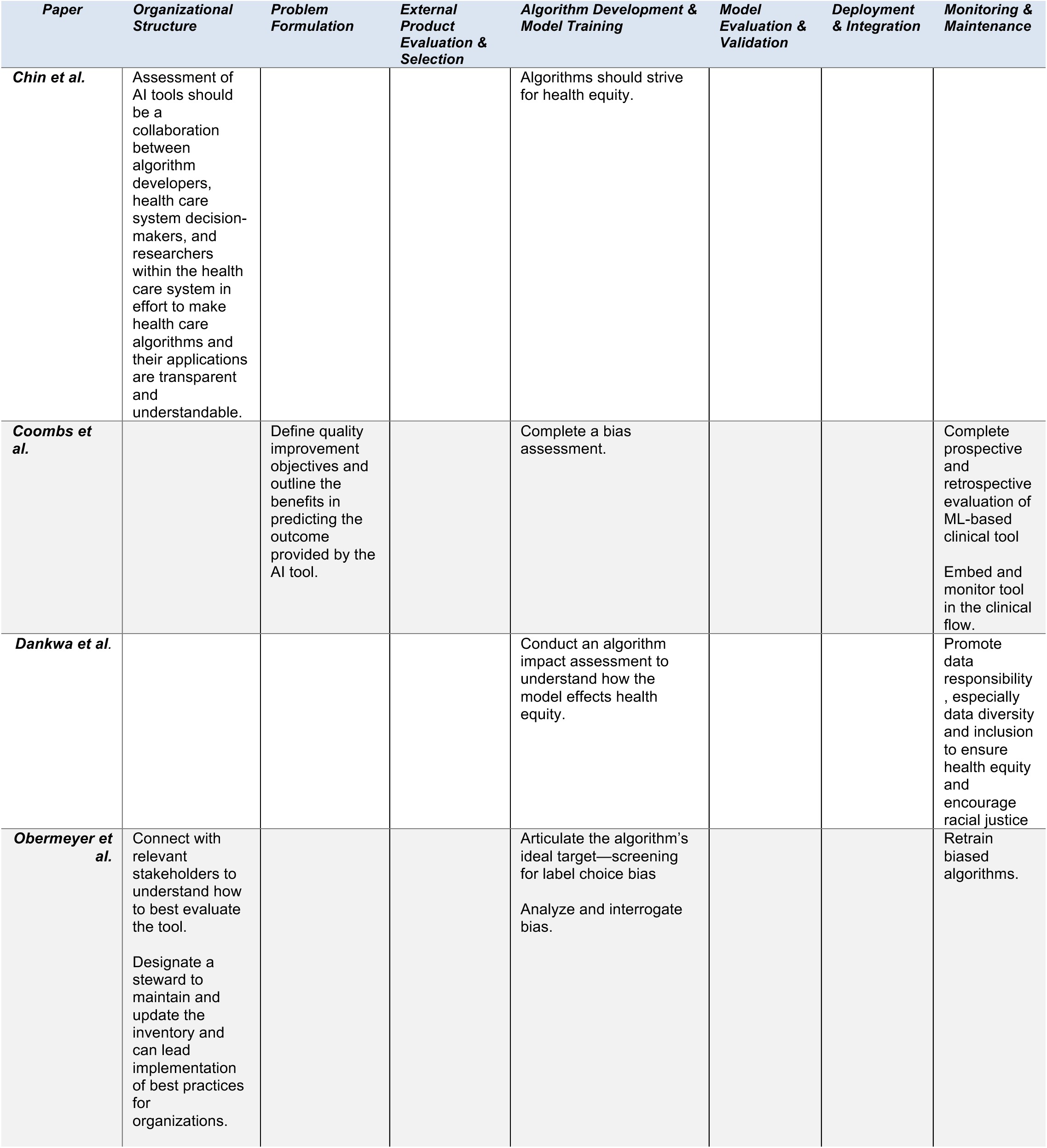
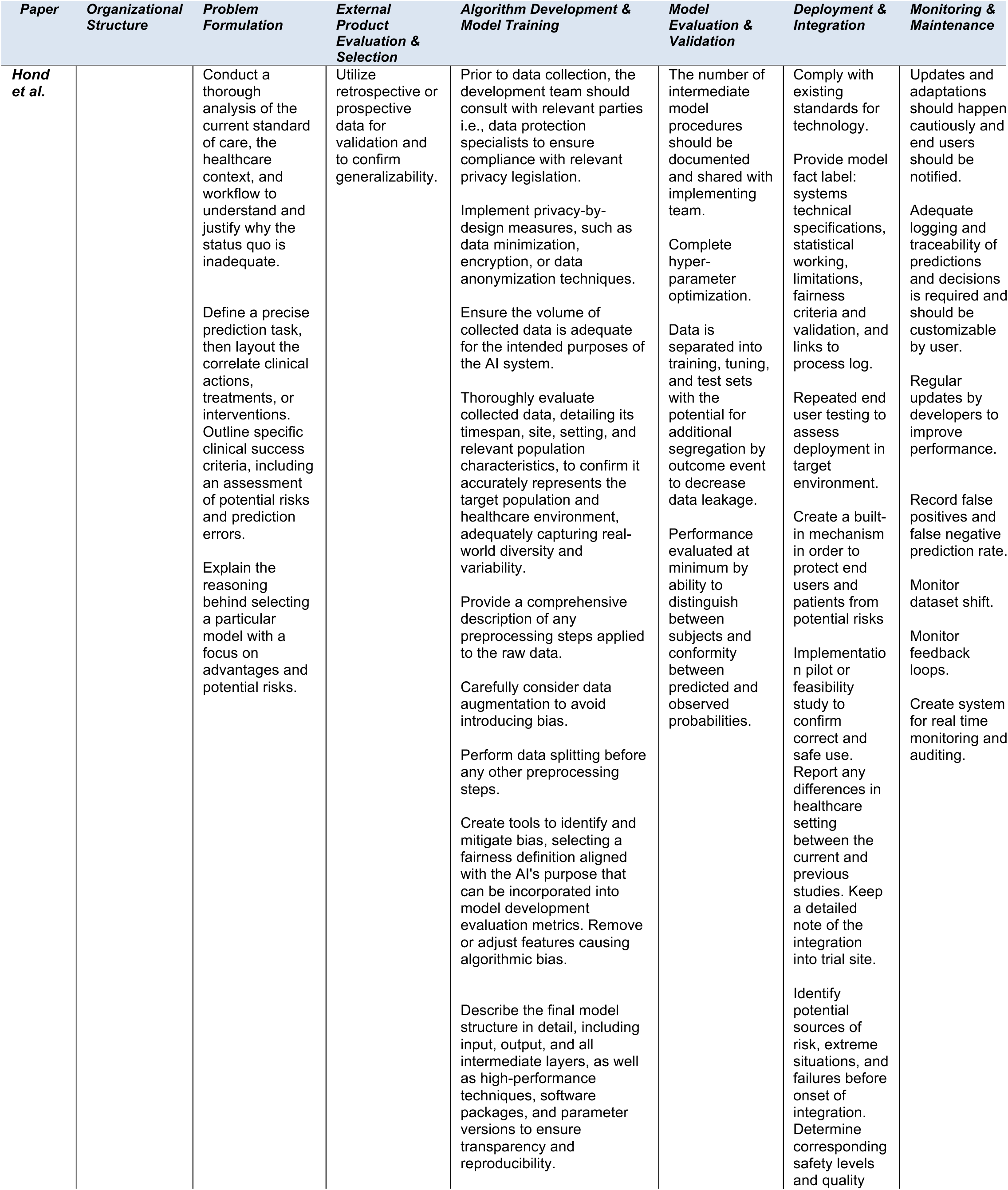
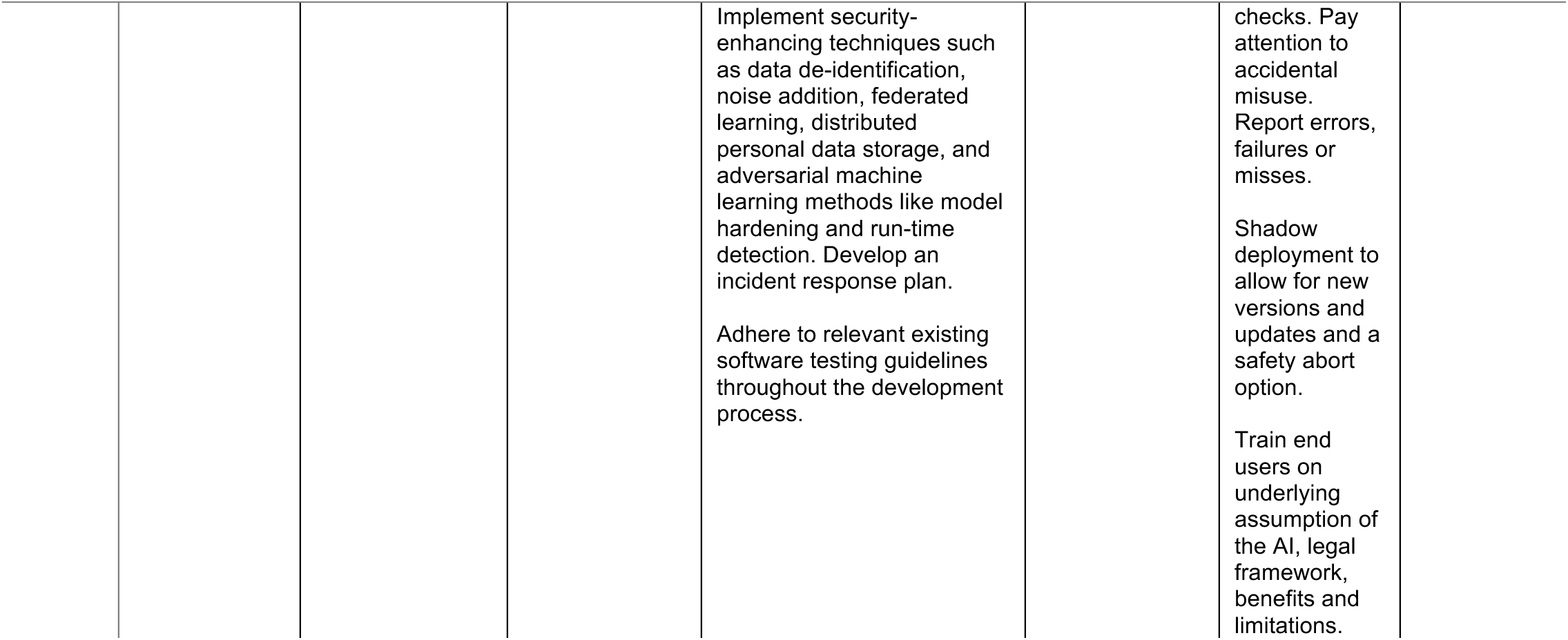
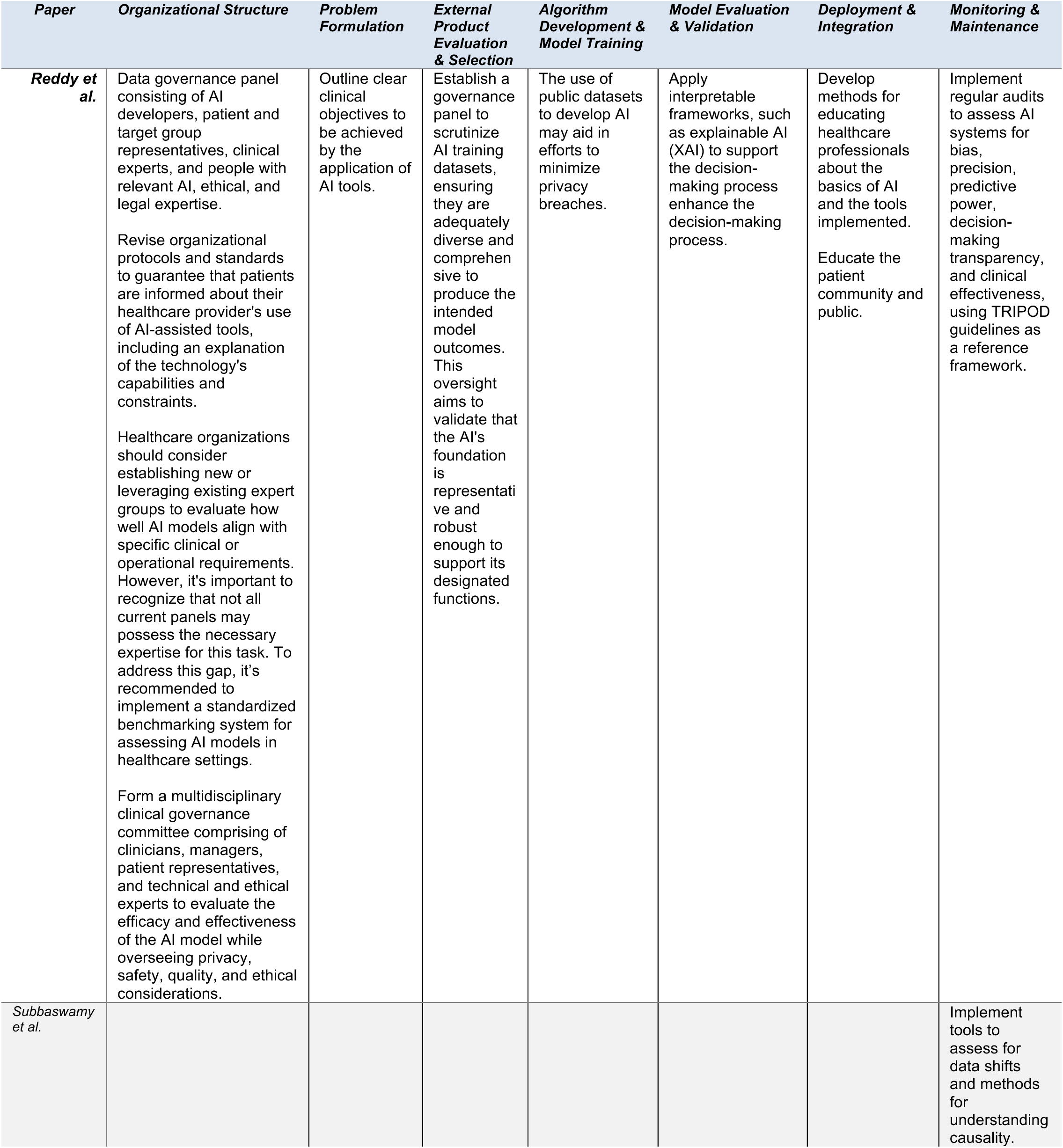
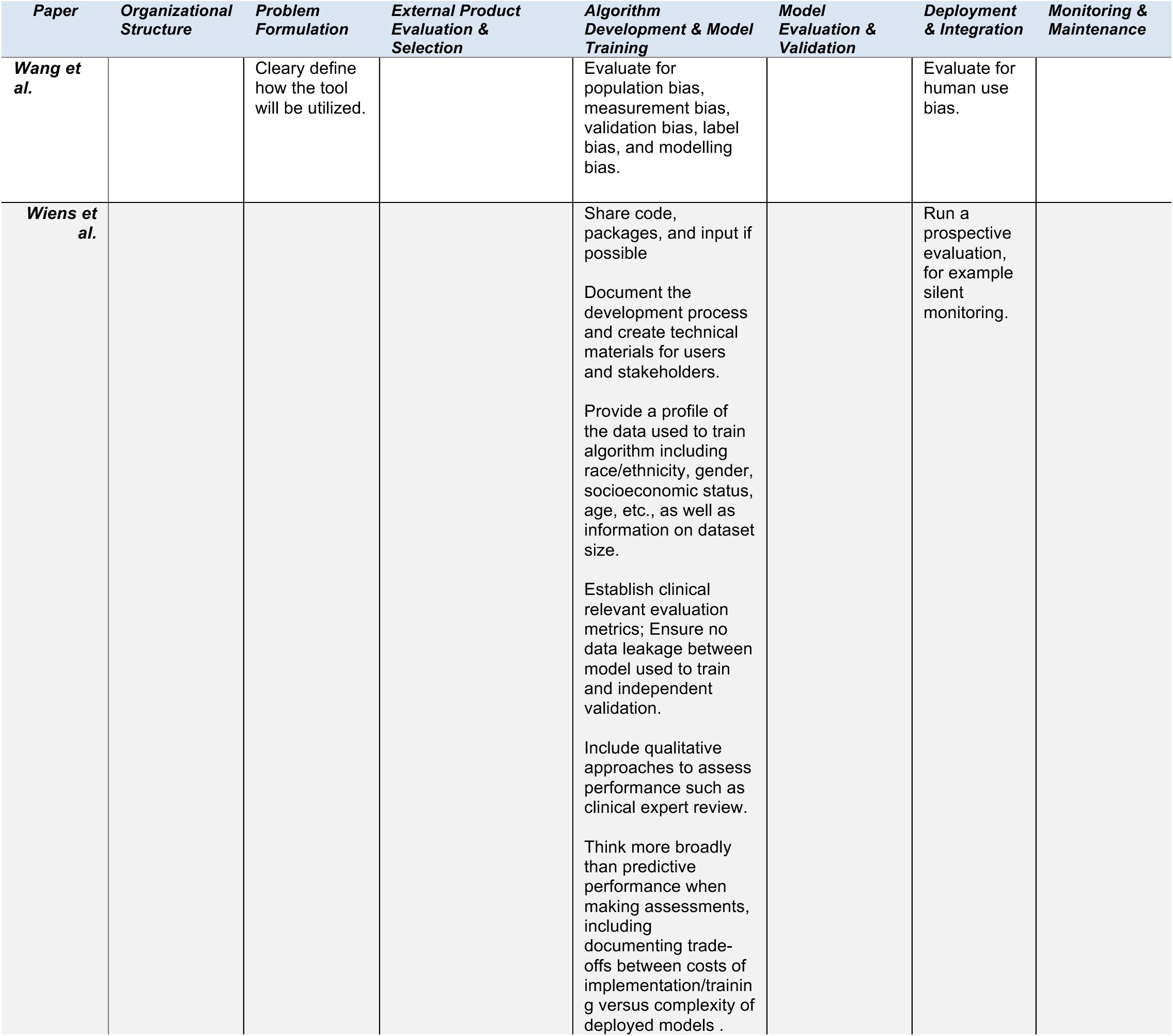
An overview of the selected publications recommendations across the different categories of the AI cycle. These are the distilled guidelines which may miss some of the details included in each articles. Four articles were excluded from this table due to the extensive level of detail in each section, i.e., checklist with a multitude of items per section.

**Supplemental Table 2.**
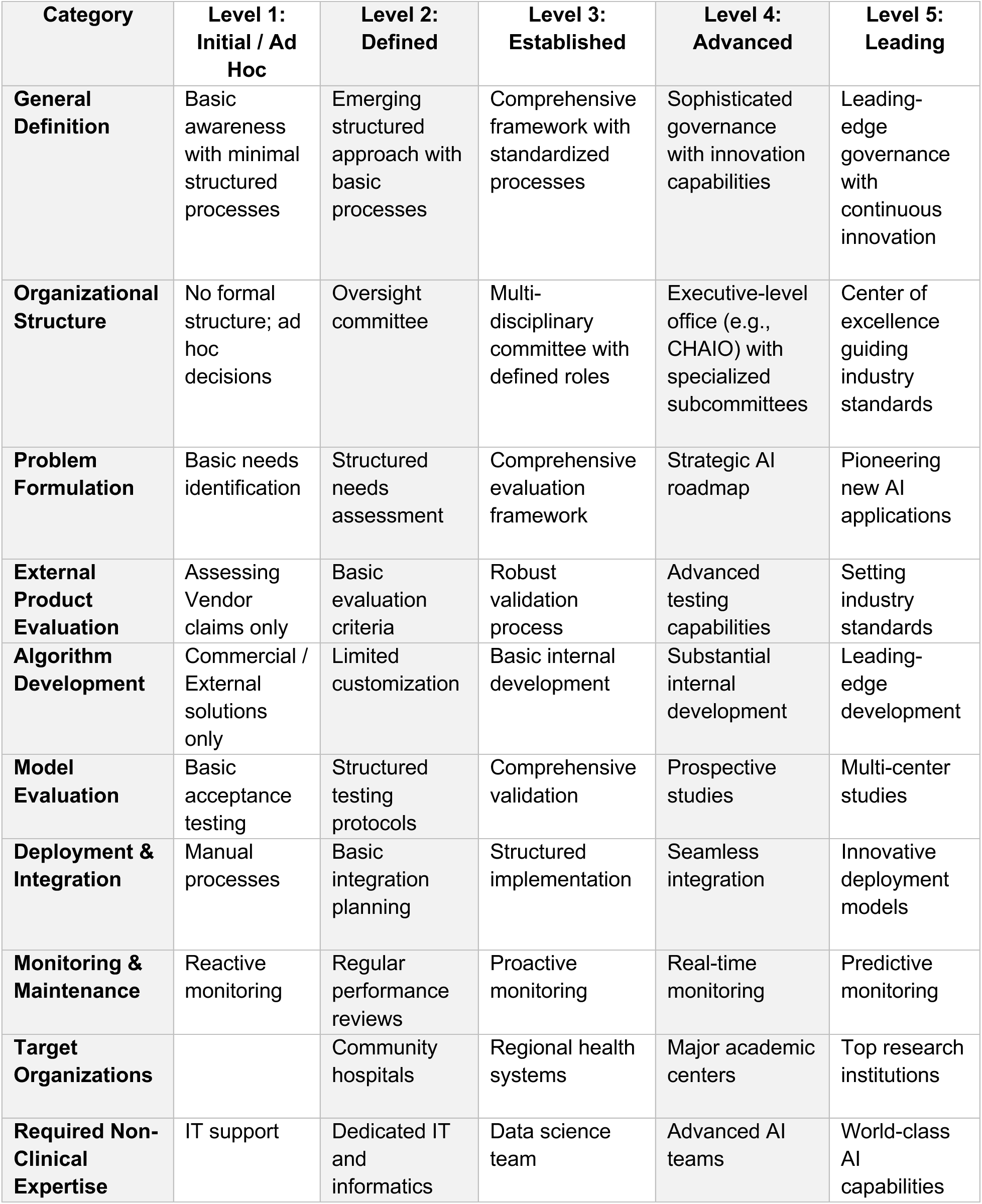
Summary version of the Healthcare AI governance Readiness Assessment (HAIRA).

| **Category** | **Level 1:**<br>**Initial / Ad**<br>**Hoc** | **Level 2:**<br>**Defined** | **Level 3:**<br>**Established** | **Level 4:**<br>**Advanced** | **Level 5:**<br>**Leading** |
| --- | --- | --- | --- | --- | --- |
| **General Definition** | Basic awareness with minimal structured processes | Emerging structured approach with basic processes | Comprehensive framework with standardized processes | Sophisticated governance with innovation capabilities | Leading-edge governance with continuous innovation |
| **Organizational Structure** | No formal structure; ad hoc decisions | Oversight committee | Multi-disciplinary committee with defined roles | Executive-level office (e.g., CHAIO) with specialized subcommittees | Center of excellence guiding industry standards |
| **Problem Formulation** | Basic needs identification | Structured needs assessment | Comprehensive evaluation framework | Strategic AI roadmap | Pioneering new AI applications |
| **External Product Evaluation** | Assessing Vendor claims only | Basic evaluation criteria | Robust validation process | Advanced testing capabilities | Setting industry standards |
| **Algorithm Development** | Commercial / External solutions only | Limited customization | Basic internal development | Substantial internal development | Leading-edge development |
| **Model Evaluation** | Basic acceptance testing | Structured testing protocols | Comprehensive validation | Prospective studies | Multi-center studies |
| **Deployment & Integration** | Manual processes | Basic integration planning | Structured implementation | Seamless integration | Innovative deployment models |
| **Monitoring & Maintenance** | Reactive monitoring | Regular performance reviews | Proactive monitoring | Real-time monitoring | Predictive monitoring |
| **Target Organizations** |  | Community hospitals | Regional health systems | Major academic centers | Top research institutions |
| **Required Non-Clinical Expertise** | IT support | Dedicated IT and informatics | Data science team | Advanced AI teams | World-class AI capabilities |

**Supplemental Table 3.** List of Journals Included in Literature Review.

|  | Journal Title |
| --- | --- |
| 1 | New England Journal of Medicine Artificial Intelligence |
| 2 | Artificial Intelligence in Medicine |
| 3 | International Journal of Medical Informatics |
| 4 | Radiology: Artificial Intelligence |
| 5 | Journal of Medical Internet Research |
| 6 | Nature Partner Journals Digital Medicine |
| 7 | IEEE Journal of Biomedical and Health Informatics |
| 8 | The Lancet Digital Health |
| 9 | Journal of the American Medical Informatics Association |
| 10 | Journal of Medical Internet Research Artificial Intelligence |
| 11 | Intelligent Based Medicine |
| 12 | BMC Bioinformatics |
| 13 | IEEE Transactions on Pattern Analysis and Machine Intelligence |
| 14 | Nature Machine Intelligence |
| 15 | New England Journal of Medicine |
| 16 | The Lancet |
| 17 | Journal of the American Medical Association |
| 18 | British Medical Journal |
| 19 | Nature Medicine |
| 20 | Nature Biotechnology |
| 21 | Nature Communications |
| 22 | Nature Computational Science |

